# Failure detection in biomedical image classification under realistic distribution shifts: insights from a large-scale evaluation

**DOI:** 10.64898/2026.05.04.26350496

**Authors:** Paul Steinmetz, Frédérique Frouin, Vincent Morard, Irène Buvat

## Abstract

Biomedical images (BMI) exhibit variability due to different acquisition protocols, devices, and patient populations, making failure detection at inference time essential for reliable deployment of clinical classifiers. As existing evaluations of failure detection methods use different settings, it is difficult to compare results and identify the best strategy, if any. We present a comprehensive evaluation of eight confidence scoring functions and two score-aggregation strategies across eight BMI tasks spanning diverse modalities, backbone architectures, training setups, and failure sources. The confidence ranking ability and classification error mitigation are jointly evaluated. While no single method systematically dominated across settings, aggregation of confidence scores consistently matched or approached the best individual method and substantially reduced silent failure rate. The failure detection performance was strongly correlated with classifier accuracy for all tested settings. These findings provide large-scale evidence of the strengths and limitations of confidence scoring strategies and offer actionable guidance for mitigating silent failures under realistic distribution shifts in BMI.

**Graphical Abstract:** 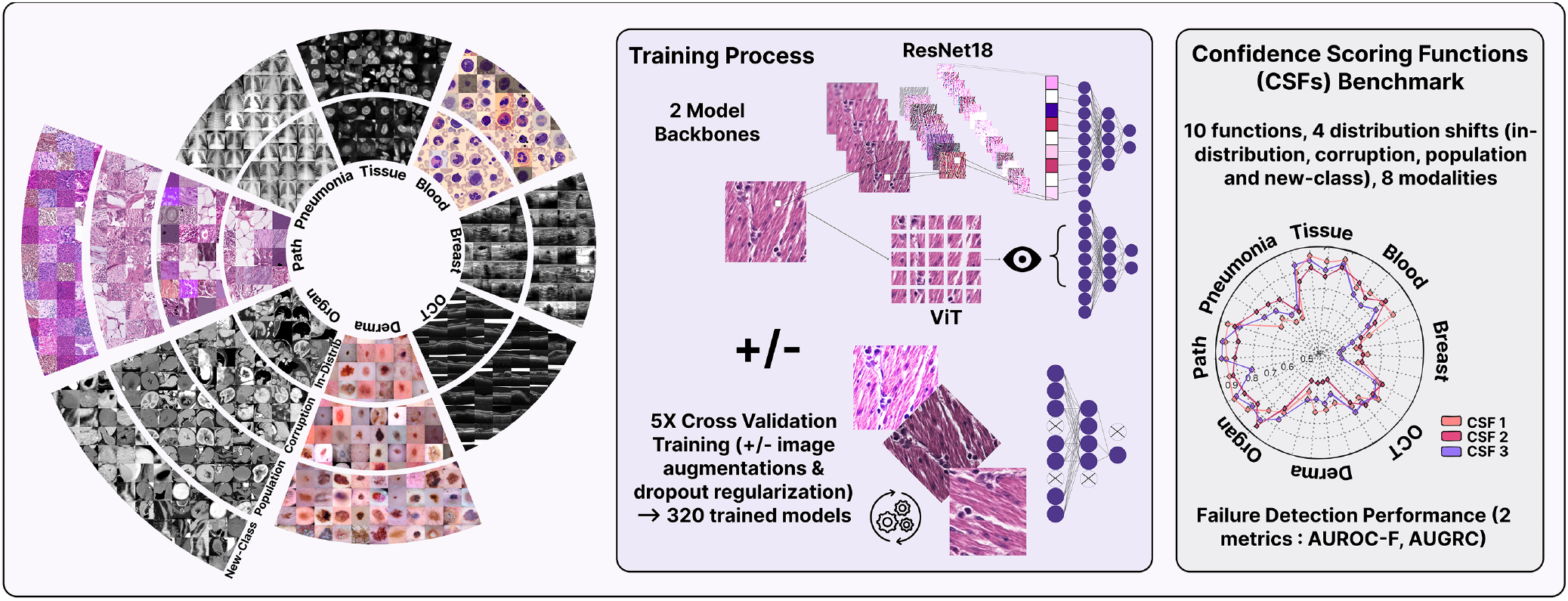

**Highlights:**

- Failure detection performance is independent of biomedical image modality but strongly correlated with classifier accuracy.
- Monte Carlo Dropout does not outperform simple margin-based scores despite substantially higher computational costs.
- Aggregating standardized confidence scores provides more robust performance than individual methods and reduces mean silent failure rates from 24% down to 8% for realistic distribution shifts.
- Cross-validation ensembling improves both classification accuracy and failure detection performance.

## 1. Introduction

With the rapid expansion of artificial intelligence across a wide range of domains, failure detection has received increasing attention over the past decade [6, 13]. This is especially true in image classification, where deep learning models have become ubiquitous. The inherent complexity of such models makes the decision-making process difficult to understand and verify (hence the so-called “black-box” terminology). This lack of interpretability is often accompanied by overconfident predictions made by the model [15]. To avoid such undesired behavior in high-stakes tasks such as medical imaging, silent failure detection methods have been developed, i.e., filtering out erroneous predictions according to an uncertainty score derived from a confidence scoring function (CSF). Related research areas, including misclassification detection, out-of-distribution (OOD) detection, uncertainty quantification, and selective classification, are now largely studied. Although they all share the same ultimate goal of mitigating model failures, they differ in their formal definitions and assumptions, making direct comparison challenging. Consequently, standardizing evaluation appears as critical as developing new methods [21].

Failure detection methods can be divided into intrinsic and post-hoc approaches. Intrinsic methods integrate uncertainty estimation into the network architecture or training process, producing uncertainty scores as part of the model output. Post-hoc methods, in contrast, operate on an already trained model. Despite the greater number of intrinsic approaches [29, 20], post-hoc methods are often easier to implement and more flexible, as they are model-agnostic (no retraining protocols or model architecture modifications are needed).

Recurrent pitfalls persist in the evaluation of failure detection methods [21]. Although real-world medical imaging involves domain changes due to different acquisition protocols, scanner hardware, patient populations, or imaging artifacts [27], failure detection methods are often evaluated on a single test dataset or under poorly characterized distribution changes. Evaluating robustness under meaningful and semantically well-defined shifts (label-preserving: covariate shifts, or non-preserving: semantic/context new-class shifts [45, 40]) is therefore essential. There is also recurring confusion in the literature between OOD and failure detection: the former aims to detect samples coming from a distribution distinct from that of the training data, whereas the latter focuses on detecting failures. Treating OOD detection as a proxy for failure detection is misleading and should be avoided: OOD data may still be correctly classified, and in-distribution (ID) misclassifications are not rare. Finally, many studies only focus on a CSF’s ability to detect failure samples, independently of the underlying classification results, while intrinsic methods, by modifying the model structure, can deteriorate classification performance.

According to recent evaluation guidelines [21], failure detection performance evaluation should be (1) reported using a unified metric reflecting both failure prediction ability and classification performance; (2) evaluated on diverse and realistic failure sources; (3) assessed through a metric quantifying the failure detection abilities of a given classifier-CSF pair, rather than through surrogate tasks such as OOD detection.

As summarized in Fig. 1, we present a large-scale evaluation of existing methods for failure detection across eight medical imaging classification tasks, using publicly available datasets that span diverse imaging modalities and classification tasks on both classical convolutional ResNet18 networks (R18) and vision transformer (ViT) families, trained under four setups (with and without data augmentation and dropout regularization). We evaluate the robustness of the scoring functions under distribution shifts by assessing both failure detection performance: AUROC-F, the area under the ROC curve for separating failures from successes and failure prevention capabilities: AUGRC, the area under the generalized risk-coverage curve, corresponding to the mean proportion of silent failures over all rejection thresholds and jointly capturing classification accuracy and failure detection performance). We thus provide a comprehensive evaluation of failure detection methods in medical imaging and assess the value of aggregation strategies.

**Figure 1.**
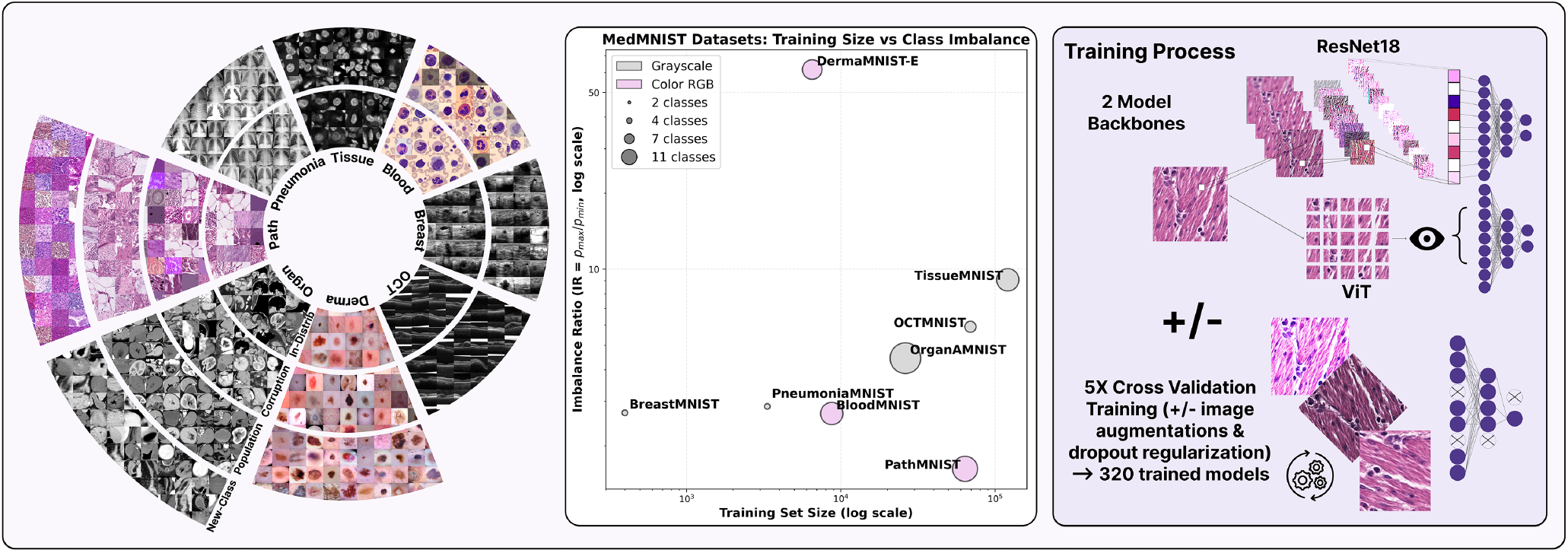
Image processing and training pipeline: 8 MedMNIST datasets used and 4 applied shifts (left) for failure detection evaluation (In-Distribution, Corruption, Population and New-Class shifts). Training dataset size and class imbalance (center). For each MedMNIST dataset, 2 backbones trained (ViT and R18) under 4 different setups (+/− data augmentation and dropout regularization) with 5-fold cross-validation training (right).

## 2. Methods

### 2.1. Datasets and Data Splits

To cover a wide range of biomedical imaging modalities and classification tasks, we selected eight datasets from the MedMNIST v2 collection [43, 12] for model training and ID evaluation. MedMNIST provides standardized 224 × 224 biomedical image datasets spanning X-ray, histology, ultrasound, computed tomography, optical coherence tomography, electron microscopy, and dermatoscopy, with both binary and multi-class classification tasks. To evaluate robustness under realistic distribution shifts, we further included four independent external datasets: DermaMNIST-E [1], HMU-CRC-Hist550K [39], MIDOG++ [4], and AMOS-22 [22], which were used as external test sets for skin lesion, colorectal histopathology, mitosis detection, and abdominal CT evaluations, respectively (Fig. 1)

For each dataset, the predefined ID test split was used for evaluation. As several CSFs require calibration, the predefined training and validation sets were merged and split into a study (80%) and a calibration (20%) set, stratified by class. The study set was then split five times into training (80%) and validation (20%) sets, stratified by class, for cross-validation. For OrganAMNIST, where multiple slices originate from the same 3-D volume, the predefined validation set was used as calibration set, and the predefined training set was split into training and validation sets while ensuring that all slices from a given volume remained in the same split. This protocol ensured strict separation between training, calibration, and evaluation data. The complete description of used dataset can be found in Appendix A.

### 2.2. Model Training and Classification Metrics

Two backbone architectures were trained for each dataset: a R18 convolutional neural network and a ViT with 16 × 16 patches, both initialized with `IMAGENET1K_V1` weights. R18 was selected as it provides competitive performance in the MedMNIST v2 benchmark [43], while ViT was included to assess the effect of transformer-based representations, which have shown strong performance in medical imaging tasks [17].

Both backbones were trained with an Adam optimizer using an initial learning rate of 10^−3^ for R18 and 10^−4^ for ViT. Binary cross-entropy was used for binary tasks and cross-entropy for multi-class tasks, with a batch size of 128.

Four training setups were evaluated: standard training (S), data augmentation (DA), dropout regularization (DO), and data augmentation with dropout (DADO). Data augmentation was performed using RandAugment [10] with two moderate-severity transformations sampled from the default transformation pool. Dropout was added after each residual block and before the fully connected layer for R18 (p=0.3), and in the attention and multilayer perceptron blocks for ViT (p=0.1). Early stopping was applied based on validation loss with a patience of 10 epochs. Overall, eight model ensembles were trained per dataset, corresponding to two backbones, two augmentation settings, and two dropout settings.

Classification performance was reported using balanced accuracy (bAcc) and area under the receiver operating characteristic curve (AUC). For multi-class tasks, AUC was computed in a one-vs-all setting and averaged over classes. Predictions were averaged across the five cross-validation folds.

### 2.3. Distribution Shifts and Failure Sources

CSFs were evaluated across three categories of failure. First, ID misclassifications corresponded to erroneous predictions on samples drawn from the training distribution. Second, covariate shifts corresponded to label-preserving shifts, where images remained assignable to the training label space but differed in their acquisition or image characteristics, including both synthetic image corruptions (CS) and population shifts (PS). Third, new-class shifts (NCS) corresponded to label-non-preserving shifts, where samples belonged to classes unseen during training.

CS images were generated using the python package MedMNIST-C [11], which applies modality-specific image corruptions. Details of applied corruptions can be found in Appendix B.

PS were evaluated using external datasets acquired at clinical centers distinct from those used for training, namely DermaMNIST-E [1], HMU-CRC-Hist550K [39], and AMOS-22 [22].

NCS were evaluated using unseen classes from AMOS-22 and MIDOG++ [4]. Following recent recommendations for failure detection evaluation [21], new-class evaluation datasets were constructed by combining correctly classified ID samples (successes) with unseen-class samples (failures), thereby removing the confounding effect of ID misclassifications.

### 2.4. Confidence Scoring Functions

We evaluated three families of CSFs: margin-based scores derived from probabilities or logits, prediction-variability scores based on repeated inference, and feature-space scores based on distances in latent space. For all CSFs, larger values indicate higher predictive uncertainty.

Let *x* denote an input sample. For binary classification, *p* = *p* (*y* = 1|*x*) denotes the predicted probability of the positive class. For multi-class classification, *p*_*c*_(*x*) = *p*(*y* = *c*|*x*) denotes the predicted probability for class *c*, and *z*_*c*_ (*x*) the corresponding logit. For CSFs relying on multiple predictions, the final class prediction was obtained by averaging predictive probabilities:

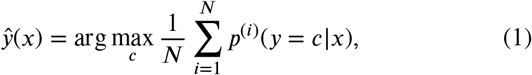

where *p*^(*i*)^(*y* = *c* | *x*) is the probability obtained from the *i*-th inference.

#### 2.4.1. Margin-Based Scores

For binary tasks, uncertainty was quantified using the distance to hard labels (DHL):

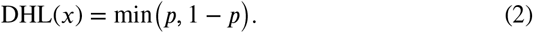

For multi-class tasks, the maximum softmax response (MSR) [19] was used:

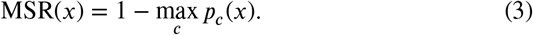

Calibrated variants were obtained using Platt scaling for binary tasks and temperature scaling for multi-class tasks. For binary classification, the pre-sigmoid logit (z) was transformed as

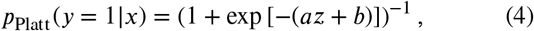

with *a* and *b* fitted on the calibration set. The calibrated score, denoted DHL-S, was then computed by replacing *p* in (2) with *p*_Platt_.

For multi-class classification, temperature scaling [15] was applied to the logits:

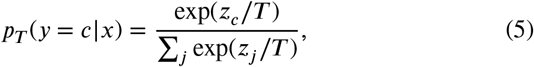

where *T* > 0 was optimized on the calibration set by minimizing the negative log-likelihood.

The maximum logit score (MLS) [18] was defined as − |*z*| for binary tasks and − max_*c*_ *z*_*c*_(*x*) for multi-class tasks.

#### 2.4.2. Prediction-Variability Scores

Deep ensembles (DE) [28] quantified uncertainty as the standard deviation of predictions across the five cross-validation models. Monte Carlo Dropout (MCD) [14] was computed by activating dropout at inference and performing N=30 stochastic forward passes. Test-time augmentation (TTA) [5] was computed from five RandAugment-transformed versions of the input image. For both MCD and TTA, uncertainty was defined as the standard deviation of the resulting predictions:

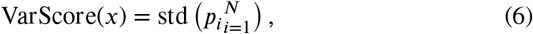

where *p*_*i*_ denotes the prediction obtained at inference *i*.

Greedy Policy Search (GPS) [32] used the calibration set to select augmentation sequences maximizing failure detection performance. At inference, the top three sequences were applied, and the final score was defined as the mean standard deviation across the selected sequences:

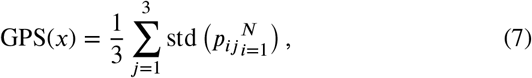

where *p*_*ij*_ denotes the prediction obtained from the *i*-th augmented sample of sequence *j*.

#### 2.4.3. Feature-Space Score

The K-nearest-neighbor score (KNN) [35] was computed in the penultimate-layer feature space. Features were first reduced using principal component analysis fitted on the training set while retaining 90% of the explained variance. The uncertainty score was then defined as

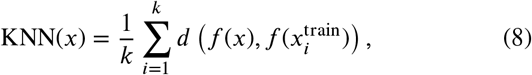

where *f* (*x*) is the latent representation of *x, d* is the Euclidean distance, and 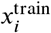 is the *i*-th nearest training sample. The value of *k* was selected on the calibration set by grid search over *k* ∈ {1, …, 200}.

### 2.5. Score Aggregation

Two aggregation strategies were evaluated. In the per-fold setting (Mean-Agg), each CSF except DE was computed independently for each cross-validation fold. For each CSF, the calibration-set mean 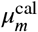 and standard deviation 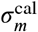 were used to standardize scores before averaging:

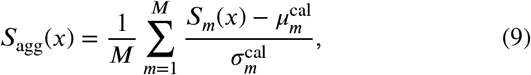

where *S*_*m*_ denotes the *m*-th CSF.

In the ensemble setting (Mean-Agg+Ens), scores were first averaged across the five folds when applicable, and then aggregated using (9). This setting included DE, KNN, GPS, MSR or DHL, MSR-S or DHL-S, MLS, and MCD when dropout layers were available. Standardization ensured comparable scales across heterogeneous CSFs.

### 2.6. Benchmark Evaluation Metrics

Failure detection was evaluated using the area under the receiver operating characteristic curve for failure detection (AUROC-F), which measures the ability of a CSF to separate incorrect from correct predictions across all thresholds.

To jointly account for confidence ranking and classification performance, we also used the area under the generalized risk-coverage curve (AUGRC) [36]. AUGRC averages the proportion of silent failures over all rejection thresholds, with lower values indicating better performance. Its closed-form expression is

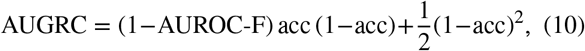

where acc denotes the baseline classification accuracy. A random CSF (AUROC-F = 0.5) yields 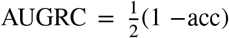, whereas an oracle CSF (AUROC-F = 1) achieves 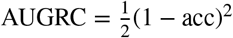.

## 3. Results

### 3.1. Model Classification Performance

Classification results on test datasets are summarized in Figure 2. Results are reported using bAcc across all datasets for both backbones and different setups.

**Figure 2.**
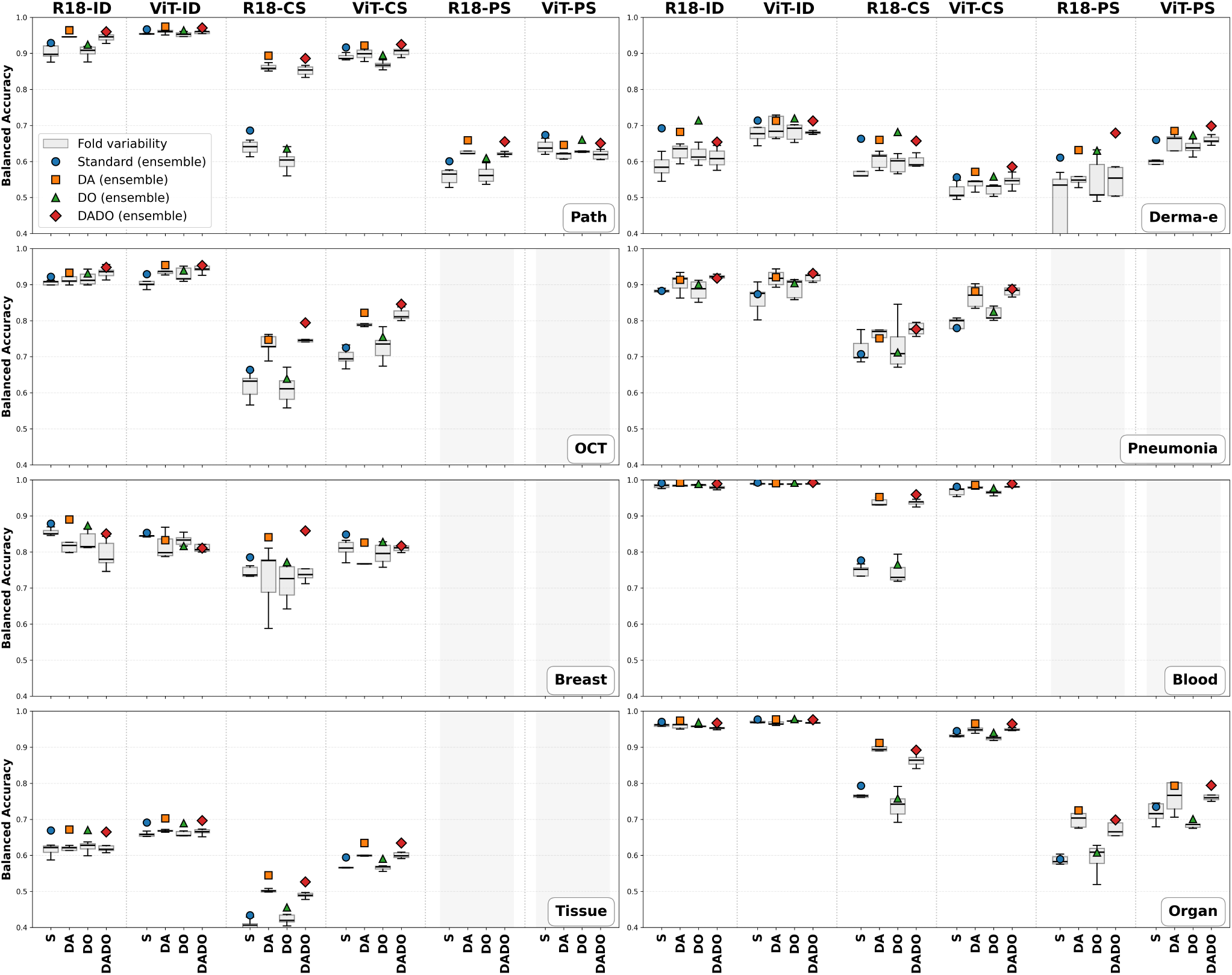
Balanced-Accuracy results (ResNet18: R18, Vision Transformer: ViT, S: Standard, DA: Data Augmentation, DO: Dropout Layers, DADO: DA+DO) across all test datasets and failure sources (ID: In-Distribution, CS: Corruption Shifts, PS: Population Shifts). Box-plots: fold results, Markers: model ensembling results. Non-applicable configurations are grayed.

The classification task difficulty varied from very simple (ID OrganAMNIST and BloodMNIST), with nearly perfect bAcc values, to more difficult tasks, particularly where datasets are imbalanced, such as TissueMNIST and DermaMNIST-E. Performance generally decreased under CS and PS evaluations; a mean drop of 0.27 in bAcc was even observed when OrganAMNIST models were evaluated on the AMOS-22 and of 0.32 when PathMNIST models were evaluated on the HMU-CRC-Hist550K PS test sets in comparison with ID test sets. In most cases, ViT achieved results comparable to or better than R18, which is consistent with prior observations [17]. BreastMNIST ID and DermaMNIST-E CS evaluations were the only two configurations where the transformer under-performed. Mean bAcc gains of 0.02, 0.09, and 0.08 were observed for ViT compared with R18 under ID, CS, and PS evaluations, respectively. While data augmentation during training almost always improved performance (0.01, 0.07, and 0.05 bAcc gains, respectively), this was not observed with the addition of dropout layers, resulting in similar results (0, 0, and 0.01 bAcc change, respectively) compared to standard training. Ensembling significantly improved performance compared to the mean per-fold bAcc across almost all use cases (0.02, 0.03, and 0.04 bAcc gains, respectively), with a maximum gain of up to 0.13.

### 3.2. CSFs Benchmark

We evaluated all 8 CSFs combined with each fold and 2 aggregation methods using ensemble models across 168 different configurations: 2 backbones x 4 training setups x (8 ID + 8 CS + 3 PS + 2 NCS datasets). The means AUROC-F and AUGRC across the 5 folds and ensembling aggregation are reported in Fig. 3 and Fig. 4 for ID and CS evaluations, and in Fig. 5 for PS and NCS evaluations.

**Figure 3.**
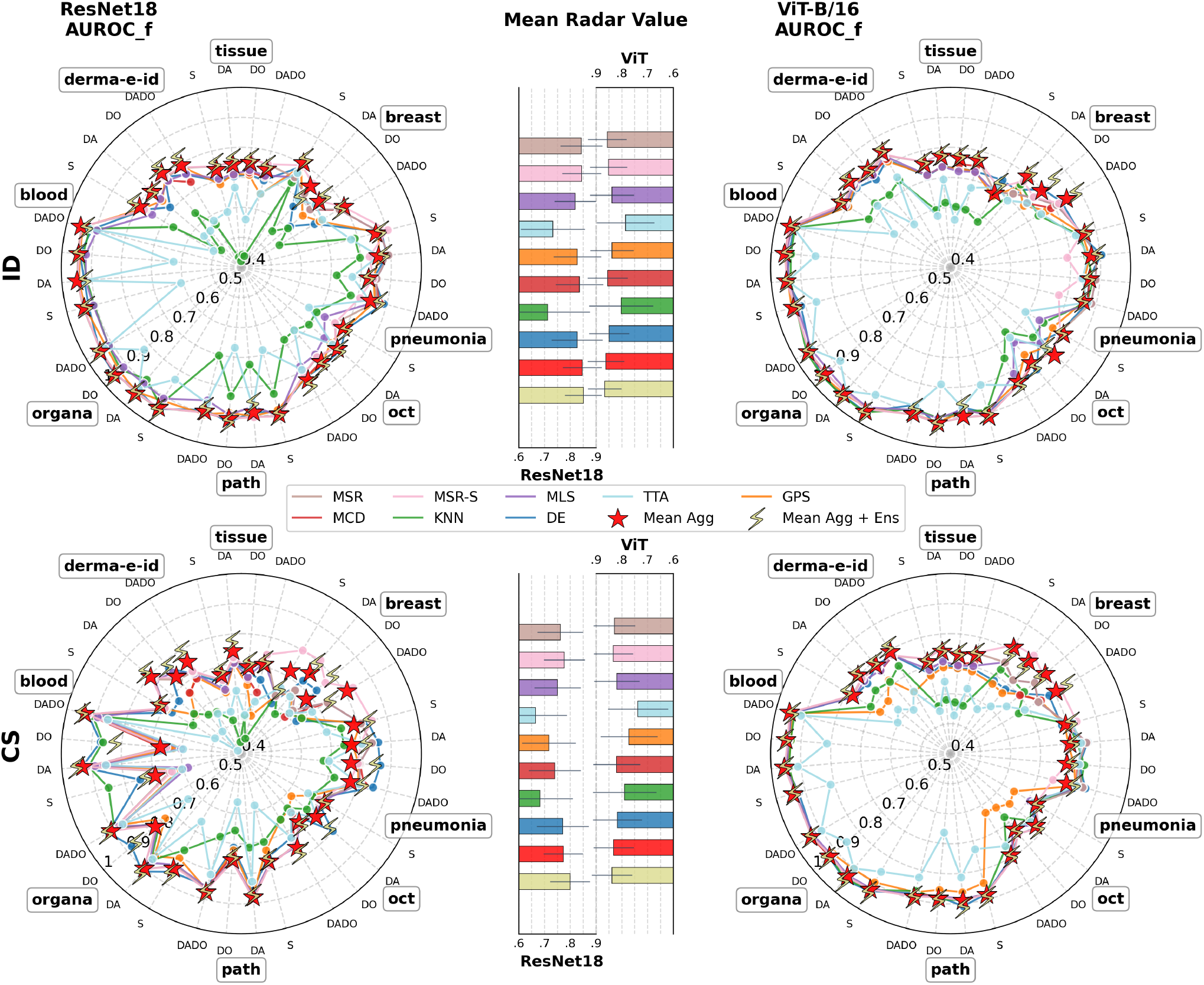
AUROC-F results across all datasets and training setups. Results are shown for two backbones (left: R18, right: ViT) under ID (first row) and CS (second row) evaluations. Mean radar-value histograms ± standard deviations across backbones and data shifts are displayed in the center.

**Figure 4.**
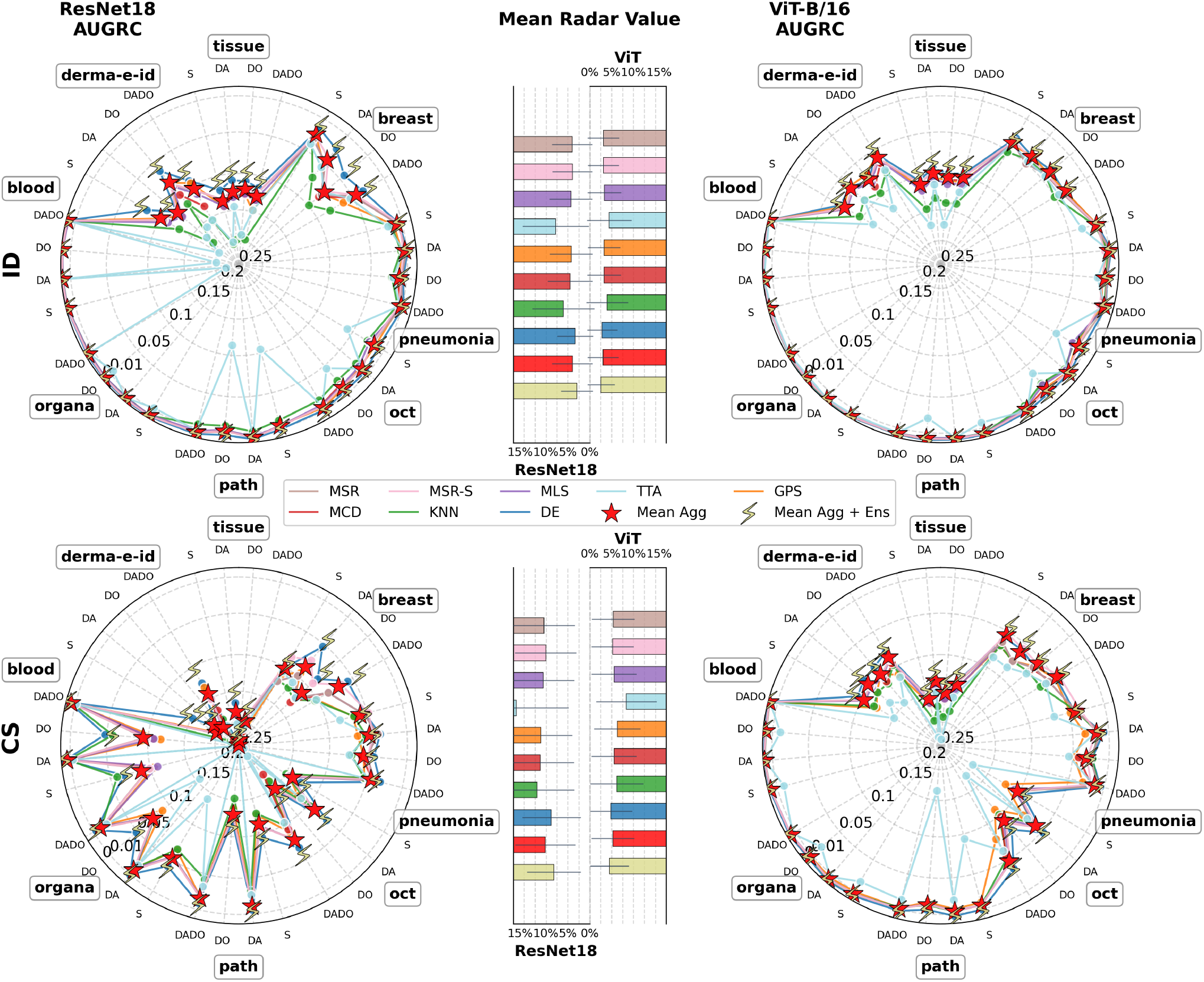
AUGRC results across all datasets and training setups. Results are shown for two backbones (left: R18, right: ViT) under ID (top) and CS (bottom) evaluations. Mean radar-value histograms ± standard deviations across backbones and data shifts are displayed in the center.

**Figure 5.**
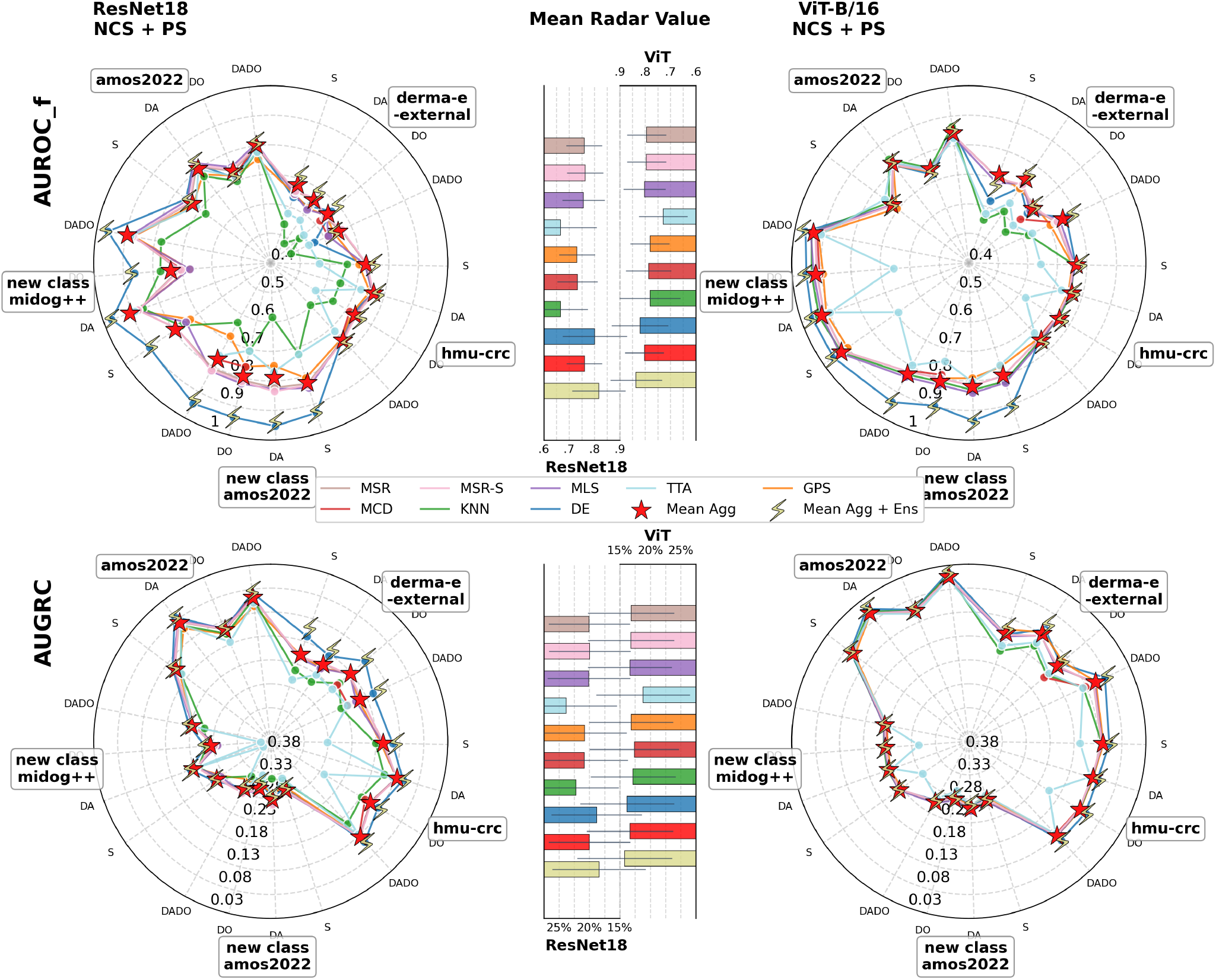
AUROC-F (top) and AUGRC (bottom) results across all datasets and training setups. Results are shown for two backbones (left: R18, right: ViT) for PS (“amos2022”, “hmu-crc”, and “derma-e-external” datasets) and NCS (“new-class amos2022”, “new-class midog++”)) evaluation. Mean radar-value histograms ± standard deviations across backbones and data shifts are displayed in the center.

#### 3.2.1. Silent failure detection performance across configurations

Regarding silent failure detection, no individual scoring function consistently outperformed the others across the evaluated experiments.

Among the lowest-performing methods, TTA exhibited strong sensitivity to the usage of data augmentation during training, oscillating between radar centers (poor failure detection) and outer regions, resulting in the smallest radar mean values across all evaluated shift types and backbones. Augmentation-based scoring methods benefited from the addition of GPS, which selected a subset of relevant corruptions prior to inference and yielded substantially more robust performance than naive TTA. This advantage somewhat vanished after noise addition. Similarly relying on multiple stochastic forward passes aggregation, MCD tended to outperform GPS; however, its performance remained below that of the DE variability scoring function under corruption shifts.

The KNN feature-based distance in latent space also showed fair to poor performance. It exhibited poorer ranking performance than other scoring functions across shifts when evaluated with the R18 backbone. In contrast, margin-based scoring functions (MSR, from scratch or after post-hoc calibration, and MLS) were more robust to the failure source nature, backbone choice, and training setup. Among these, MSR frequently achieved the highest radar mean value among individual CSFs in joint label space configuration evaluation (ID and CS).

Similar rankings were observed when analyzing AUGRC. By construction, classification performance changes for methods relying on stochastic prediction aggregation and for methods that intrinsically alter the predictive model, whereas post-hoc, stochastic CSFs share the same underlying model and differ solely in their confidence ranking behavior. While TTA, GPS and MCD failed to outperform margin-based scores, DE demonstrated higher AUGRC than the other stochastic CSFs. This is partly due to the substantial gain in classification accuracy obtained through cross-validation, which was not observed with TTA, GPS or MCD.

#### 3.2.2. Backbone and shift effects

Radar plots and mean histograms highlight the superiority of using a ViT backbone over a CNN architecture. This remained true regardless of the evaluated shift category and for almost all CSFs (*average* ± *std* best CSF AUROC-F increase of 0.01 ± 0.03, 0.03 ± 0.04, 0.02 ± 0.04, and 0.01 ± 0.04 across setups and datasets for ID, CS, PS and NCS evaluations, respectively). This was even more obvious when taking into account the classifier performance due to the overall higher bAcc of ViT over R18 (Fig. 2), especially when the test data distribution differed from the training one with increased differences in radar mean values between CS, PS, or NCS vs ID. ViT improved best-CSF AUGRC by 0 ± 1%, 4 ± 3%, 3 ± 3%, and 2 ± 1% compared with ResNet18 for ID, CS, PS, and NCS evaluations, respectively. The performance rankings of CSFs were similar for the ViT and R18 architectures.

Substantial drops in performance were observed when comparing CS or PS with ID results.

Best-performing CSFs across setups demonstrated a mean drop of 0.04 ± 0.05 (R18) and 0.03 ± 0.03 (ViT) in AUROC-F for CS evaluation. This degradation was associated with an average increase in the mean proportion of silent failures across rejection thresholds (mean AUGRC) of 5 ± 4% and 2 ± 2% for R18 and ViT, respectively. The direct comparison of DermaMNIST-E subgroup results (same centers as training vs. external images), as well as that of the CT organ images (OrganAMNIST vs. AMOS-22) and of the colorectal tissue slides (PathMNIST vs HMU-CRC-Hist550K), enables quantification of the performance gap between ID and PS. An average drop in best CSF AUROC-F of 0.15±0.03 (0.13 for cutaneous lesions, 0.17 for CT organs classification, and 0.14 for histology slides) was observed for the CNN backbone, while the experiments with the transformer showed a decrease of 0.13 ± 0.03 (0.1 and 0.16, 0.14, respectively) for PS compared to ID. This reduced performance in failure detection came along with a mean increase of 10 ± 1% (8%, 11% and 12%, respectively) and 8 ± 1% (6%, 7%, and 11%, respectively) in AUGRC with R18 and ViT, respectively.

Our results also illustrate a greater challenge in detecting failures vs. detecting OOD samples composed of unseen semantics but within the same context (near-OOD). Best CSF performance on OOD AMOS-22 CT organs detection reached a mean across training setups AUROC-F of 0.94 and 0.92 whereas predicting failures on known-label images from AMOS-22 achieved a mean performance of 0.79 and 0.8, for R18 and ViT, respectively. Accordingly, OOD detection on the dog cancer histology slides dataset showed better performance than HMU-CRC-550K PS failure detection in the colorectal tissue classification task: 0.93 vs. 0.77 for R18 and 0.97 vs. 0.77 for ViT AUROC-F on average across all best CSFs of each setup.

#### 3.2.3. Relationship between failure detection performance and classification accuracy

Correlations between the failure detection performance of the evaluated CSFs (AUROC-F) and the corresponding classifier bAcc are shown in Fig. 6. For all CSFs, a positive linear correlation was observed, with an overall mean Pearson correlation coefficient of *r* = 0.76 and regression slope coefficients ranging from 0.43 (more horizontal) to 0.71 (closer to identity function). Performance of margin-based and aggregation methods was less dependent on classification bAcc than the performance of other CSFs while KNN presented the steepest slope.

**Figure 6.**
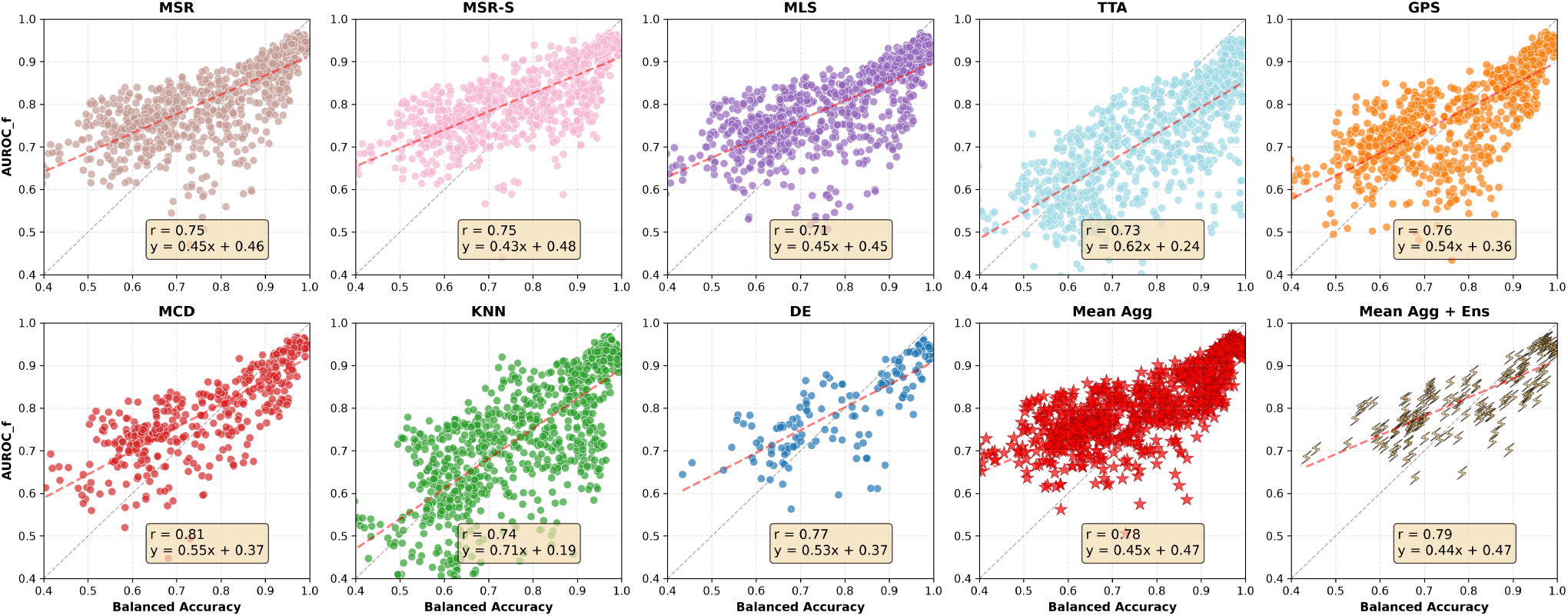
CSF AUROC-F correlation with classification bAcc (over all configurations: training setup + backbone + dataset) across failure sources

#### 3.2.4. CSF Aggregation

For all 168 evaluated configurations, the mean aggregation of multiple CSFs matched or closely approached the best-performing individual CSF, resulting in consistently higher radar mean AUROC-F and AUGRC than those of individual methods. The combination of DE response variability (blue histograms) with the aggregation of the other CSFs (red histograms) resulted in overall higher performance (yellow histograms) across all configurations (Fig. 3–Fig. 5). Fig. 7 illustrates the average reduction of silent failures proportion (failure samples retained after rejection) across all rejection thresholds for all model backbones and Mean-agg+Ens-CSF pairs compared to the original error rate with no failure prediction ability. An average reduction of 82 ± 9%, 76 ± 10%, 66 ± 5%, and 65 ± 2% respectively in ID, CS, PS and NCS was achieved thanks to the rejection mechanism of uncertain samples.

**Figure 7.**
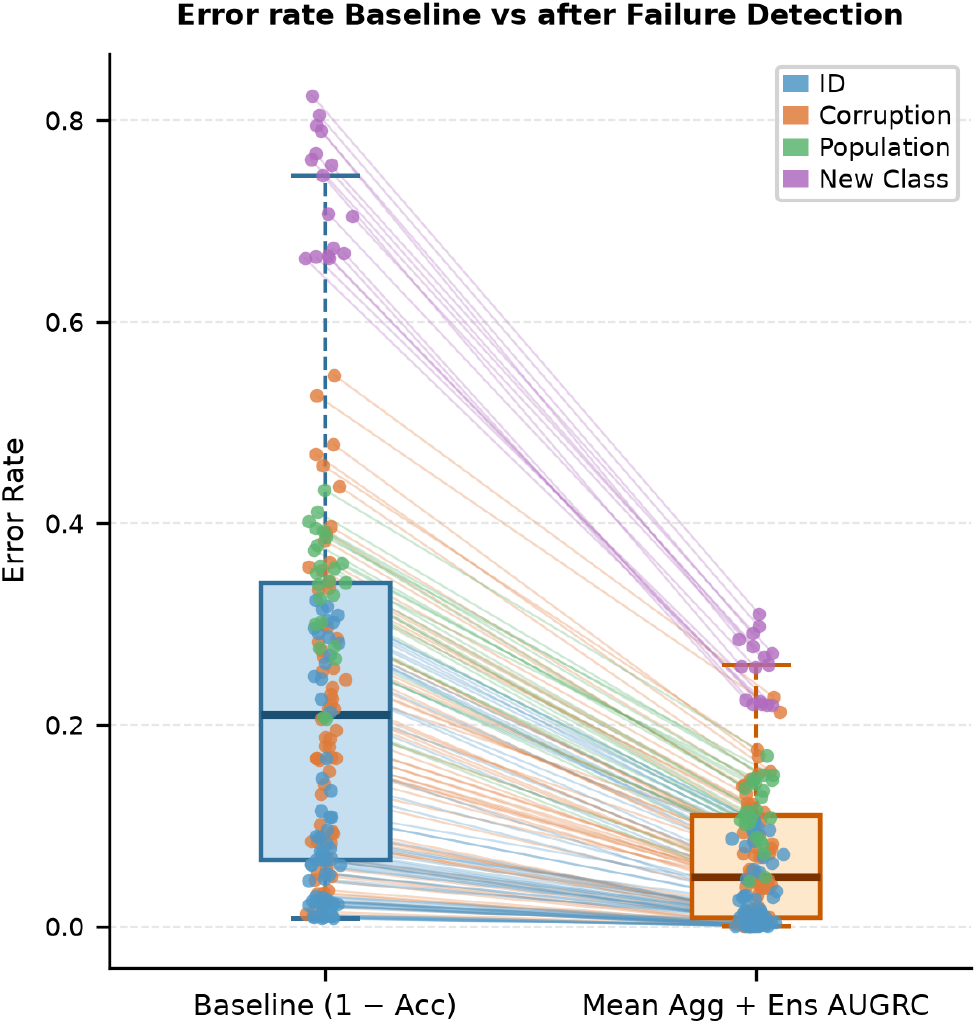
Original error rate without CSF vs AUGRC achieved with Mean-Agg+Ens across all test shifts, datasets, and training setups.

## 4. Discussion

Our extensive evaluation revealed a strong and consistent positive relationship between classifier performance and failure-detection performance across datasets, backbones, and distribution shifts (Fig. 6). This suggests that the quality of the underlying classifier is a major determinant of failure-detection performance, with poorly performing models generally yielding less reliable confidence estimates. This important relationship has never been evaluated to the best of our knowledge. This also highlights the robustness of some CSF with respect to poor baseline classification accuracy: KNN performance are highly correlated with classification performance, while the reliability of margin-based and aggregation strategies are much less dependent on the classification performance.

Although no single scoring function proved superior across all model architectures and shift types, aggregation strategies consistently achieved competitive performance across evaluation settings, which suggests potential complementary aspects of CSFs. This supports the use of aggregation strategies in challenging classification tasks. While some method combinations such as MCD-MSR (softmax output over multiple forward passes with MCD, [21]), or Ensemble+Mean predictive entropy in a segmentation benchmark (mean entropy computed from ensembling results [46]) were reported, the benefit of our original Z-score aggregation strategy of independent CSFs has not been investigated.

The discrepancy between CSFs performance ranking across failure sources has been identified in previous benchmarks [33, 8, 21, 34]. Yet our evaluation extends these observations to a larger and more diverse collection of medical imaging modalities, spanning histopathology, radiography, computed tomography, dermoscopy, optical coherence tomography, and blood-cell microscopy and included evaluation of methods that were never compared before.

Our findings also challenged several previously reported conclusions on individual methods. We evaluated two augmentation based CSFs, with either random corruption (TTA, used in 7% of reviewed studies [29]) or after optimization using a calibration hold-out set (GPS). These rarely outperformed margin-based methods. While TTA was highly sensitive to the training strategy, GPS generalized better to different training setups (with or without DA). These findings highlight the importance of carefully selecting the applied augmentations, as unknown corruption drastically perturbs failure detection. While still performing better than TTA, GPS performance was reduced under distribution shifts suggesting that the choice of calibration set is important when selecting augmentations. In particular, calibration data drawn from the same distribution as the target test set may yield better results. In contrast to our results, Yang et al. [44], in the OpenOOD benchmark found that augmentation-based methods ranked among the top three CSFs across all configurations. Similarly, in a histopathological classification use case, Mehrtens et al. [33], showed superior performance of TTA under weak and strong shifts on ID data vs MCD or MSR. Both cited studies evaluated CSFs under a single training setup, whereas our results show that training augmentations can substantially affect TTA performance.

We also demonstrated throughout our experiments that MCD, while being computationally intensive, rarely out-performed simple MSR. Yet, previous work on diabetic retinopathy detection [30] (a single dataset of 88,702 fundus images) showed that a margin-based method (entropy) performed substantially worse than MCD across data shifts. The variability in CSF performance across datasets observed in our benchmark suggests that CSF rankings may strongly depend on the target application and may explain this conflicting results. Yet, MCD is the most frequently used scoring function in the literature (38% of reviewed studies vs. 13% using MSR [29]).

In joint label space regimes (ID, CS or PS), while not always outperforming other CSFs in terms of AUROC-F, DE variability consistently achieved near-best AUGRC values. This improved ranking is driven by the higher baseline classification accuracy obtained through cross-validation ensembling compared with single-fold models (Figs. 4 and 5). Such improvements in classification performance were not observed for the other stochastic methods (MCD, TTA, and GPS). This further emphasizes the importance of accounting for classifier performance when evaluating silent-failure mitigation [21, 36]. For the NCS evaluation, DE achieved significantly better performance for the detection of OOD samples while a reduced efficiency of MSR was noted (Fig. 5, 1st row), with even greater superiority for the CNN backbone. This clear change in CSFs ranking emphasizes the differences between failure and OOD detection tasks and may reflect poorer performance of margin-driven metrics in favor of other types of scoring functions as the shift in semantics increases.

A similar degradation of margin-based methods with increasing shift severity was reported by Olivares and Brockmeier [34] for CNNs. The authors further observed superior performance of geometry-aware scoring functions based on latent-space distances. Similarly, the OpenMIBOOD benchmark [16] reported strong KNN performance alongside poor MSR results. In contrast, KNN consistently underperformed relative to other CSFs across most of the evaluated shift scenarios in our benchmark. This feature-based superiority had already been questioned previously [21], where the prevalent Mahalanobis-distance scoring function performed well for major context changes but ranked worse under nuanced covariate shifts. In fact, failure cases can strongly activate characteristics of learned class manifold and thus be projected deeply within the latent training distribution [8] which can mislead confidence estimates. In our benchmark, KNN significantly outperformed the other CSFs in only one setting: blood-cell classification with a standard R18 model under corruption shifts. This may indicate a stronger alteration of image characteristics than in the other evaluated scenarios. These discrepancies highlight the limited transferability of conclusions across dataset settings and should encourage caution regarding the widespread use of feature-based approaches for failure and OOD detection.

While some recent work on natural images has attempted to bridge the gap between failure detection and OOD detection by combining MSR for failure detection and KNN for OOD detection into a nonlinear fusion score [42], its applicability to biomedical imaging remains uncertain given the limited performance of KNN observed in our benchmark. More generally, some of these conflicting results with the literature may arise from differences between natural and medical images and may reflect the impossibility of relying on results obtained when evaluating CSFs on natural images. This was demonstrated by Gutbrod et al. [16], who showed that the CSFs performing best on natural images often performed poorly on biomedical images due to significant differences in image features and context.

Our results illustrated an important mean reduction in silent failure rate across all rejection thresholds (Fig. 7). Yet, the addition of a CSF does not completely prevent silent failures, with more than 20% of failures remaining undetected in some use cases. The inability to reject all failures has previously been used to argue against implementing CSFs in clinical routine [8]. Our results challenge that claim: while confidence scoring functions do not allow for perfect mitigation, the relative robustness to training shifts and classifier performance of the aggregated score (combining at least DE variability with MSR scores) might be sufficient for some applications. The relationship between classifier and failure detection performance should be taken into account when interpreting confidence estimates in practice, with more caution in CSF results when the classifier performs poorly. Such deferral strategies have already been evaluated in practical clinical settings for breast cancer and tuberculosis screening, where MSR is incorporated into a probabilistic framework to determine when cases should be deferred to expert review [13]. The authors report a 5–15% reduction in false positives compared to AI-only systems at matched false-negative rates.

Our systematic analysis provides a broad and systematic evaluation of silent failure prevention methods; yet several limitations should be noted. Only 8 CSFs were evaluated, while many other scoring functions have been proposed in the literature. This is particularly true in the OOD detection research field, where numerous feature-based approaches are described. We selected KNN over the more classical Mahalanobis distance because it does not assume that the training-data embedding follows a Gaussian distribution. Better performance of KNN relative to Mahalanobis distance has also been reported previously [35]. Furthermore, these 8 tested methods are common and easily implementable. Another limitation is that to implement such a failure detection module in practice, a holdout set should be used to infer a threshold for decision rejection. We used AUGRC metric instead, averaging the results after rejection across all possible thresholds. We believe that with an appropriate calibration set, results after rejection at an optimized threshold should outperform reported AUGRCs. Shift magnitudes (CS, PS or NCS) were not quantified, which prevented a breakdown analysis of CSF performance as a function of distribution shift importance. However, the global positive relationship with model accuracy in ID, CS and PS, alongside the overall good OOD detection performance in NCS, suggest a higher sensitivity to learned embeddings than to shift magnitudes or image modality. Only synthetic corruptions were applied to evaluate CSFs under corruption shifts rather than using true noisy images. This, however, allowed controlled noise addition relevant to the evaluated images modality. It also made direct comparison of ID misclassifications and CS evaluation possible, as the original images used under both regimes are the same. A further limitation is that all experiments were conducted on datasets resized to a common resolution of 224 × 224 pixels. Consequently, some fine-grained image details present in native clinical images may have been lost, potentially affecting performance. Nevertheless, image down-sampling to fixed input dimensions is a standard component of most deep-learning pipelines, and the use of standardized image resolutions further enabled controlled comparisons across modalities, architectures, and distribution shifts.

## 5. Conclusion

Reliable biomedical image classification requires not only high accuracy but also mechanisms to prevent silent failures under the distribution changes that routinely occur in practice. We performed an extensive comparison of eight common failure detection methods in medical imaging across eight tasks, multiple realistic failure sources, and two backbone families with four training setups.

Across 168 configurations, we showed that no CSF consistently outperformed the others. Yet, aggregation of scores mitigated the shortcomings of individual methods across specific settings. Margin-based scores remained robust under distribution shifts within the training label space, whereas DE variability was more effective under semantic changes.

We identified a strong correlation between failure detection performance and classifier performance, highlighting the importance of accounting for the quality of the learned embedding when relying on CSF. Overall, our benchmark comprehensively describes the performance of failure detection strategies and demonstrates that appropriate score aggregation can substantially reduce the average proportion of silent failures across rejection thresholds.

## Data Availability

Images used are publicly available at:
https://medmnist.com/
https://amos22.grand-challenge.org/
https://midog2021.grand-challenge.org/
https://www.nature.com/articles/s41597-026-06675-9#Sec7
preprocessed data are available on HuggingFace: https://huggingface.co/datasets/pstnmz/FailCatcher-datasets

https://amos22.grand-challenge.org/

https://midog2021.grand-challenge.org/

https://huggingface.co/datasets/pstnmz/FailCatcher-datasets

https://www.nature.com/articles/s41597-026-06675-9#Sec7

## Data Availability

All datasets used in this study are publicly available. The MedMNIST datasets were obtained from the MedMNIST collection [43, 12]. Additional datasets include DermaMNIST-E [1], the AMOS22 dataset from the Abdominal Multi-Organ Segmentation Challenge [22], the HMU-CRC-Hist550K [39], and the MIDOG++ dataset [4].

The AMOS22, HMU-CRC-Hist550K, and MIDOG++ datasets were further preprocessed to match the input format used in our experiments. The preprocessing scripts and dataset splits required to reproduce these processed datasets are publicly available in the accompanying code repository. The trained MedMNIST models are available at: https://huggingface.co/pstnmz/FailCatcher-models

The preprocessed AMOS22, HMU-CRC-Hist550K and MIDOG++ datasets are available at: https://huggingface.co/datasets/pstnmz/FailCatcher-datasets

## Code Availability

All code required to reproduce the benchmark is publicly available at: https://github.com/pstnmz/FailCatcher

The repository includes: (i) preprocessing scripts used to convert the AMOS22, HMU-CRC-Hist550K, and MIDOG++ datasets into the input format used in this study; (ii) the full benchmarking pipeline for model training and evaluation of confidence scoring methods; and (iii) the confidence scoring toolbox developed in this work and used to compute all evaluated uncertainty scores.

## Declaration of Generative AI in Scientific Writing

During the preparation of this manuscript, the authors used generative AI tools (ChatGPT, OpenAI) to assist with language editing and clarity improvement, and GitHub Copilot (Claude Sonnet) to support code development. The authors reviewed, validated, and edited all outputs generated by these tools and take full responsibility for the content of the manuscript and the implementation of the methods.

## CRediT authorship contribution statement

**Paul Steinmetz:** Conceptualization, Methodology, Software, Investigation, Formal analysis, Visualization, Writing – Original draft. **Frédérique Frouin:** Methodology, Supervision, Writing – Review & Editing. **Vincent Morard:** Methodology, Writing – Review & Editing. **Irène Buvat:** Conceptualization, Supervision, Funding acquisition, Writing – Review & Editing.

## A. Dataset details

**Table 1.**
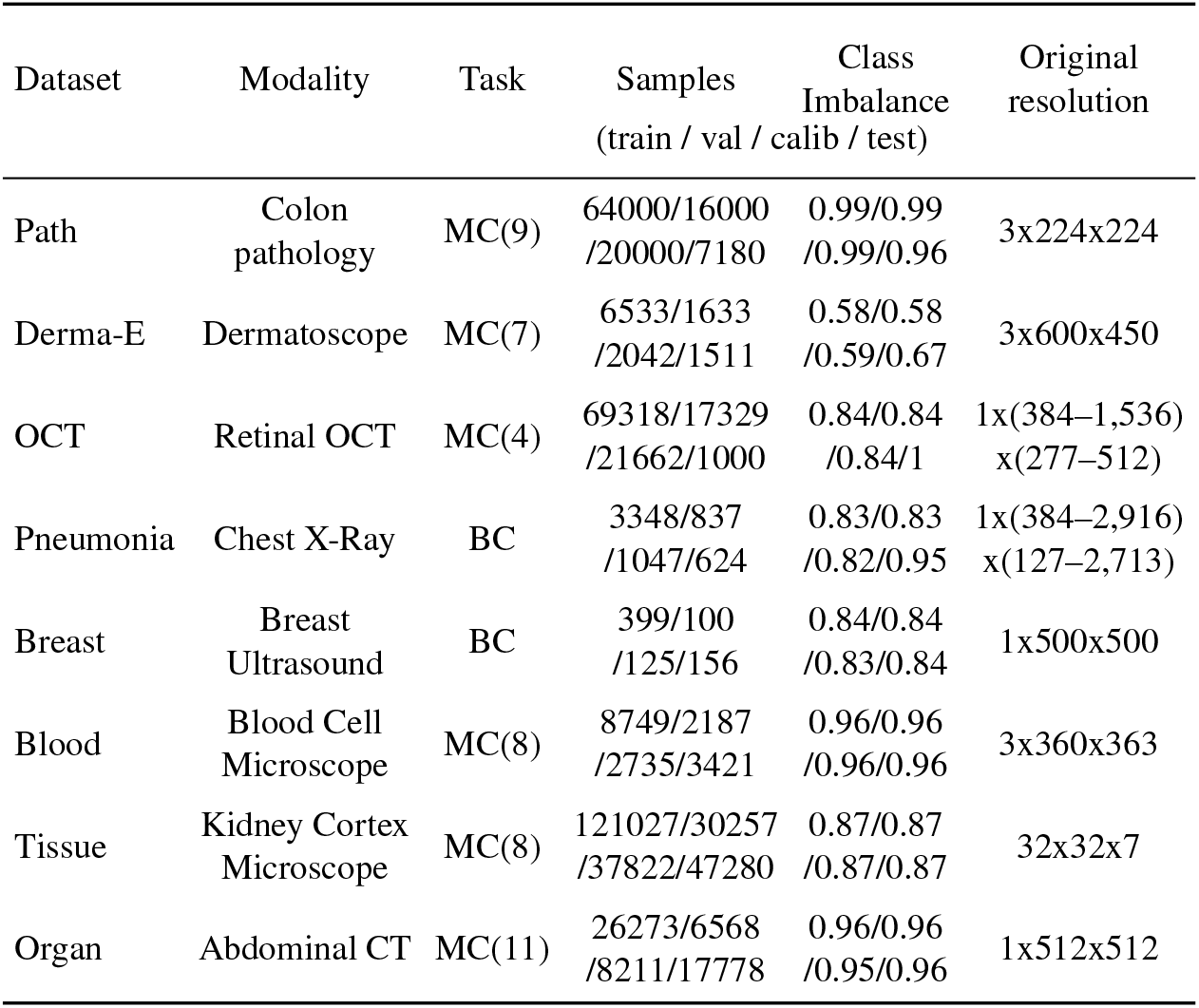
MedMNIST datasets details. Class Imbalance measured with Shannon’s Equitability (ranging from 0 for total imbalance to 1 for perfect balance)

**Table 2.** Summary of datasets used in this study. BC: Binary Classification, MC: Multi-Class Classification.

| Dataset | Description |
| --- | --- |
| **OCT** | OCTMNIST is composed of 109,309 optical coherence tomography (OCT) images of the retina from 4,686 patients [26, 25]. The task is 4-class classification of retinal conditions: choroidal neovascularization, diabetic macular edema, drusen, and normal. |
| **Derma-E** | We used the extended dermatoscopic lesion pictures DermaMNIST-E [1], utilizing the complete HAM10000 dataset [38, 37] for training and additional ISIC 2018 images [9] for validation and testing. The task is 7-class skin lesion classification. |
| **Pneumonia** | PneumoniaMNIST is based on 5,856 pediatric chest X-ray images [26, 25]. Binary classification between normal and pneumonia. |
| **Breast** | BreastMNIST consists of 780 mammography images from 600 patients [3]. Binary classification between normal/benign and malignant. |
| **Blood** | BloodMNIST contains 17,092 images of individual normal blood cells [2]. 8-class classification of blood cell types. |
| **Tissue** | TissueMNIST contains 236,386 human kidney cortex cell images drawn from BBBC051 [41, 31]. 8-class kidney cell classification. 32x32 images are upsampled to a size of 224x224. |
| **OrganA** | OrganAMNIST includes 58,850 axial CT images of 11 abdominal organs from LiTS [7]. Multi-class organ classification. |
| **AMOS-22** | AMOS-22 [22] is used as an external test set for OrganAMNIST. After preprocessing, 4,454 CT slices were obtained, including overlapping and unseen organs used for near-OOD evaluation. |
| **Path** | PathMNIST [23, 24] contains 100,000 training and 7,180 test colorectal histology patches. 9-class tissue classification. |
| **MIDOG++** | MIDOG++ [4] contains 503 histological whole-slide images. The canine subset was used as new-class shift data after patch extraction (12,450 samples). |
| **HMU-CRC-Hist550K** | HMU-CRC-Hist550K [39] contains 550,000 annotated image tiles derived from 500 whole-slide images, fully labeled into eight distinct TME tissue classes, matching PathMNIST classes. |

## B. Covariate shifts description

**Table 3.** MedMNIST-C corruption types and shift evaluations (ID: In-Distribution, CS: Corruption Shifts, PS: Population Shifts, NCS: New-Class Shifts, NA:Not Applicable)

| Dataset | Shift Type | Corruption Types |
| --- | --- | --- |
| BloodMNIST | ID, CS | brightness, bubble, contrast, defocus blur, JPEG compression, motion blur, pixelate, saturate, stain deposit |
| BreastMNIST | ID, CS | brightness, contrast, JPEG compression, motion blur, pixelate, speckle noise |
| DermaMNIST-E | ID, CS | black corner, brightness, characters, contrast, defocus blur, Gaussian noise, impulse noise, JPEG compression, motion blur, pixelate, shot noise, speckle noise, zoom blur |
| OCTMNIST | ID, CS | contrast, defocus blur, JPEG compression, motion blur, pixelate, speckle noise |
| OrganAMNIST | ID, CS | brightness, contrast, gamma correction, Gaussian blur, Gaussian noise, impulse noise, JPEG compression, pixelate, shot noise, speckle noise |
| PneumoniaMNIST | ID, CS | brightness, contrast, gamma correction, Gaussian blur, Gaussian noise, impulse noise, JPEG compression, pixelate, shot noise, speckle noise |
| TissueMNIST | ID, CS | brightness, contrast, Gaussian blur, impulse noise, JPEG compression, pixelate |
| PathMNIST | ID, CS | brightness, bubble, contrast, defocus blur, JPEG compression, motion blur, pixelate, saturate, stain deposit |
| AMOS-22 | PS, NCS | NA |
| DermaMNIST-E-external | PS | NA |
| HMU-CRC-Hist550K | PS | NA |
| MIDOG++ | NCS | NA |

